# Time to reinfection and vaccine breakthrough SARS-CoV-2 infections: a retrospective cohort study

**DOI:** 10.1101/2022.02.07.22270613

**Authors:** Sevda Molani, Andrew M. Baumgartner, Yeon Mi Hwang, Venkata R. Duvvuri, Jason D. Goldman, Jennifer J. Hadlock

## Abstract

**Background:** It is important to understand how BNT162b2, mRNA-1273, and JNJ-78436735 COVID-19 vaccines, as well as prior infection, protect against breakthrough cases and reinfections. Real world evidence on acquired immunity from vaccines, and from SARS-CoV-2 infection, can help public health decision-makers understand disease dynamics and viral escape to inform resource allocation for curbing the spread of pandemic.

**Methods:** This retrospective cohort study presents demographic information, survival functions, and probability distributions for 2,627,914 patients who received recommended doses of COVID-19 vaccines, and 63,691 patients who had a prior COVID-19 infection. In addition, patients receiving different vaccines were matched by age, sex, ethnic group, state of residency, and the quarter of the year in 2021 the COVID-19 vaccine was completed, to support survival analysis on pairwise matched cohorts.

**Findings:** Each of the three vaccines and infection-induced immunity all showed a high probability of survival against breakthrough or reinfection cases (mRNA-1273: 0.997, BNT162b2: 0.997, JNJ-78436735: 0.992, previous infection: 0.965 at 180 days). The incidence rate of reinfection among those unvaccinated and previously infected was higher than that of breakthrough among the vaccinated population (reinfection: 0.9%; breakthrough:0.4%). In addition, 280 vaccinated patients died (0.01% all-cause mortality) within 21 days of the last vaccine dose, and 5898 (3.1 %) died within 21 days of a positive COVID-19 test.

**Conclusions:** Despite a gradual decline in vaccine-induced and infection-induced immunity, both acquired immunities were highly effective in preventing breakthrough and reinfection. In addition, for unvaccinated patients with COVID-19, those who did not die within 90 days of their initial infection (9565 deaths, 5.0% all-cause mortality rate), had a comparable asymptotic pattern of breakthrough infection as those who acquired immunity from a vaccine. Overall, the risks associated with COVID-19 infection are far greater than the marginal advantages of immunity acquired by prior infection.

## Introduction

As of November 5, 2021, more than 192 million people in the United States have been fully vaccinated against severe acute respiratory syndrome coronavirus 2 (SARS-CoV-2), the virus which causes novel Coronavirus 2019 (COVID-19) (1). Vaccination against COVID-19 with the mRNA vaccines, BNT162b2 (Pfizer-BioNTech) and mRNA-1273 (Moderna), and the adenoviral vectored vaccine, JNJ-78436735 (Janssen) has significantly reduced the incidence of COVID-19 infection and associated severe outcomes (2). Centers for Disease Control and Prevention (CDC) reported the risk of infection, hospitalization and death rates among unvaccinated persons were 4.6, 10.4, and 11.3 times higher compared to those of fully vaccinated persons during the era where the Delta variant was predominant (2). Studies to date suggest that COVID-19 vaccines authorized for use in the United States (US) protect against most COVID-19 variants in the US (2). However, with the spread of the Delta variant (B.1.617.2), vaccine protection was reduced and breakthrough infections, hospitalization, and death increased among vaccine recipients (3–6). In addition to vaccine-induced immunity, the infection-induced immunity acquired from natural COVID-19 infection has shown a high effectiveness in preventing reinfection (7). According to the World Health Organization (WHO), most people who have recovered from COVID-19 develop a strong protective immune response which remains for at least 6 to 8 months after infection (8).

Previous studies that examined vaccine and infection-induced immunity in preventing COVID-19 infection have established the effectiveness of vaccines in preventing new infections, however, they are limited with small sample size, short study period interval, and insufficient consideration of spatiotemporal variables (9–15). Here, we address this gap and examine COVID-19 infection post-vaccination in the western US from December 12, 2020 to November 5, 2021, encompassing the emergence and dominance of newer variants including Delta. Additionally, we examine reinfection among unvaccinated patients to investigate the infection-induced immunity.

## Methods

This study was a retrospective cohort study on electronic health record (EHR) data from Providence-St. Joseph Health (PSJH). PSJH is a community health system with 51 hospitals and 1085 clinics across five states in the western US: Alaska, California, Montana, Oregon and Washington. In this study, we defined two cohorts, vaccine- and infection-induced cohorts (Figure 1). The vaccine-induced cohort period was from 12/12/2020 to 11/05/2021, and the infection-induced cohort period was from the beginning of the pandemic to 11/05/2021. The source population was patient with at least one medical encounter during the study period (N=7,620,084). Our outcome of interests were breakthrough and reinfection for vaccine- and infection-induced cohorts, respectively. These were defined based on the positive SARS-CoV-2 PCR test result.

**Figure 1.**
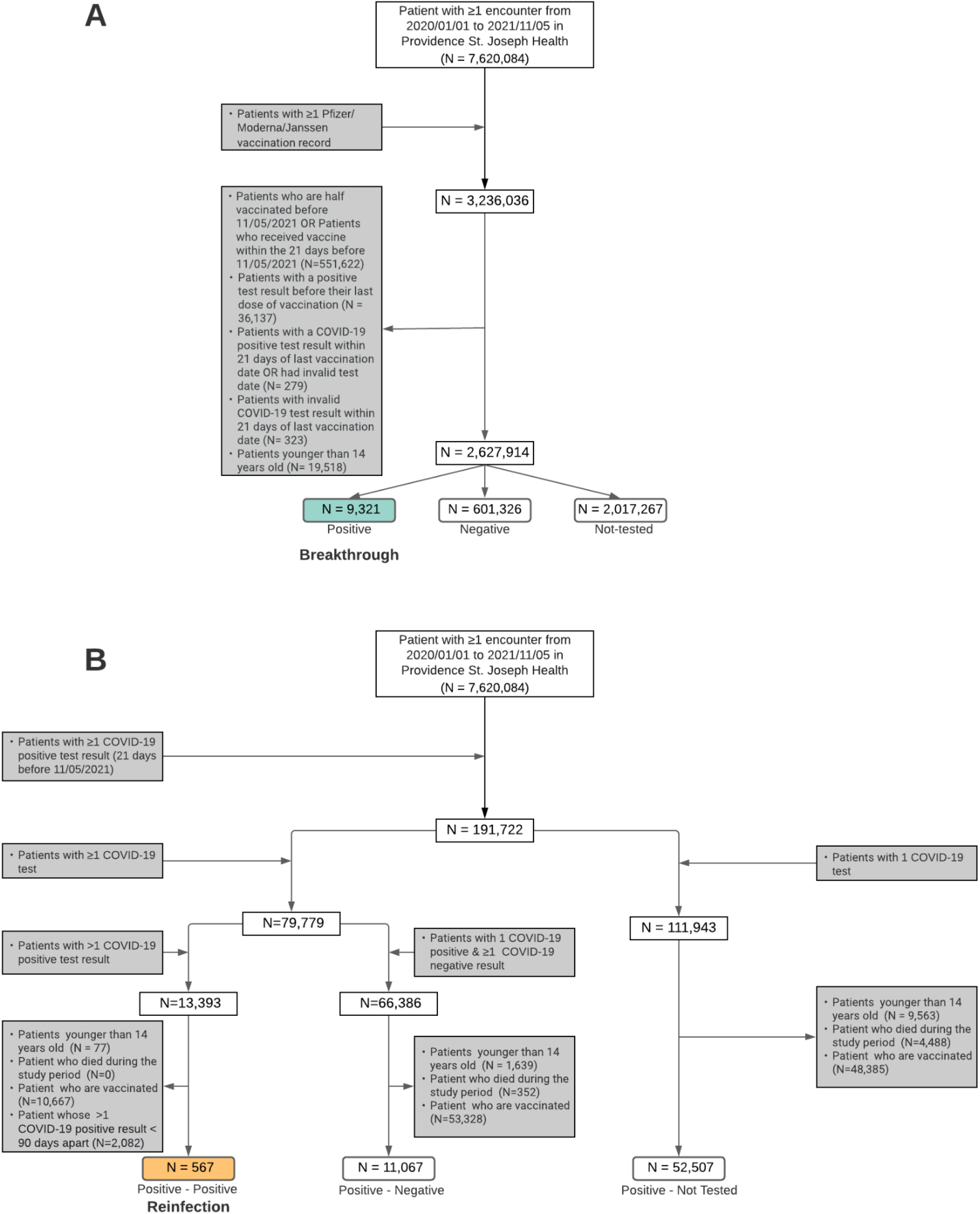
(A) Vaccine-induced immunity cohort, N=9,321 persons experienced vaccine breakthrough infection and (B) Infection-induced immunity cohort from PSJH-EHR data, N=567 reinfection cases occurred.

For the vaccine-induced immunity cohort (Figure 1A), we included patients vaccinated with at least two doses of BNT162b2 and mRNA-1273, and one dose of JNJ-78436735 during the study period (N=3,236,036). Patients who received boosters or died after their last dose of vaccine were right-censored. We excluded patients who were not fully vaccinated 21 days before 11/05/2021, had a positive test for COVID-19 before or within 21 days from the last dose of vaccination, or who received invalid test results. We also excluded patients younger than 14 years old. From this population (N=2,627,217), we categorized patients into three groups. The vaccine-induced immunity cohort was defined by SARS-CoV-2 PCR positive test after the recommended initial doses of COVID-19 vaccine: one for JNJ-78436735 and two for BNT162b2 and mRNA-1273 (N=9,321). We identified the negative group as the patient who received only negative results (N=601,326). Patients who didn’t get tested were defined as the not-tested group (N=2,017,267).

For the infection-induced immunity cohort (Figure 1B), we included patients who received at least one positive COVID-19 during the study period (N=191,722). From this population, we split into two cohorts; 1) those with multiple COVID-19 test results (N=79,779) and 2) those with one COVID-19 test result (N=111,943). Our exclusion criteria were patients who died within 90 days of initial infection, patients younger than 14 years old, and patients vaccinated during the study period. The cohort with multiple COVID-19 tests was split again into two groups: 1) a group of patients with multiple positive test results (N=13,393) and 2) a group of patients with one positive test result (N=66,386). For both groups, we applied common exclusion criteria. One positive test result group was defined as positive-negative group (N=11,067), after exclusion. The infection-induced immunity cohort was defined as ≥1 positive SARS-CoV-2 PCR test > 90 days apart, following CDC guidelines (16,17). We identified the positive-not tested group (N=52,507) by applying common exclusion criteria to the cohort with one COVID-19 test result.

We present descriptive analyses of both cohorts as frequencies and percentage for categorical variables, and as mean and standard deviation (std) for numerical variables (Supplementary Tables 1 and 2). Medical conditions include known risk factors for poor COVID-19 outcomes reported in the literature, as well as coarse grained geographical (by state) and temporal information (quarter of the year 2021). For biomedical precision we used the Systematized Nomenclature of Medicine Clinical Terms (SNOMED–CT©) hierarchy (Supplementary Table 4). For each SNOMED code listed, all descendant SNOMED codes were included. The differences between distributions of days to breakthrough was calculated using Mann Whitney U-test. Results were considered statistically significant at a (2-tailed) p-value < 0.05.

Additionally, we compared the acquired immunity through administration of each of COVID-19 vaccines using propensity score matched survival functions. Propensity score matching is a statistical method that defines a control group by matching control and treatment group based on a distribution conditioned on a fixed set of covariates (propensity score) (18,19). These covariates include age, sex, ethnic group, state of residency, and the quarter of 2021 when last dose of COVID-19 vaccine was administered. We matched the control and treatment group 1:1 ratio with replacement using nearest neighbor matching. We use the python package scikit-learn v0.23.2 (20) to calculate and match propensity scores. Missing values for continuous variables were imputed using the median value across the cohort. We confirmed covariates were balanced across control and treatment groups using standardized mean differences (SMD). Covariates were considered balanced when standardized mean difference is below 0.10(19) .

This study was approved by the Institutional Review Board (IRB) at PSJH with Study Number STUDY2020000196. Consent was waived because disclosure of protected health information for the study was determined to involve no more than a minimal risk to the privacy of individuals. We follow STROBE reporting guidelines (Supplemental Table 5).

## Results

### Vaccine-induced immunity cohort

We present descriptive analyses of vaccinated patients in Supplementary Table 1 and Supplementary Table 2. A total of 2,627,914 patients were eligible as the vaccine-induced immunity cohort. The mean age of this cohort was 53.57 years (±19.56). This cohort was enriched with patients with female sex, non-Hispanic/Latino ethnicity, White/Caucasian race, and California residence. 1,350,422 (51.4%) were administered two doses of BNT162b2 vaccine, 1,087,796 (41.4%) were administered two doses of mRNA-1273, and 189,696 (7.2%) were administered one dose of JNJ-78436735. A majority of patients completed their initial vaccination during the second quarter of 2021 (51.4%). This cohort was categorized into three groups based on their outcome (Figure 1); 9,321 (0.355%) patients were identified as vaccine breakthrough group, 601,326 (22.88%) patients in the test-negative group, and 2,027,924 patients in the not-tested group. The vaccine breakthrough group was enriched with patients between 18 to 44 years (25.6%), females (55.7%), and patients vaccinated during the first quarter of 2021 (52.0%). Figure 2 shows the distribution of time until breakthrough. The frequency of positive cases for each vaccine has a mean (std) of 161.8 (±55.4) and 152.5 (±55.9) for mRNA-1273 and BNT162b2, respectively, with approximately normal distributions, whereas JNJ-78436735 showed a more uniform distribution with a decaying tail and a mean of 123.6 (±53.7) days (Figure 2A). In addition, Figure 2B presents the normalized frequency distribution of the breakthrough cases for each vaccine. We obtain this from Figure 2A by dividing the counts in each bin by the total number of counts multiplied by the size of the bin. Furthermore, the Mann-Whitney U test showed statistically significant differences between all three distributions with p-values <0.001 for each pairwise comparison. Figure 3 presents the estimated survival functions for the outcome of breakthrough infection. In addition, Figure 4 represents the estimated survival function for the breakthrough infection in pairwise matched cohorts using propensity score matching. Note that the flat lines in post-vaccination COVID-19 breakthrough positive test shows the absence of new positive cases after that time point.

**Figure 2.**
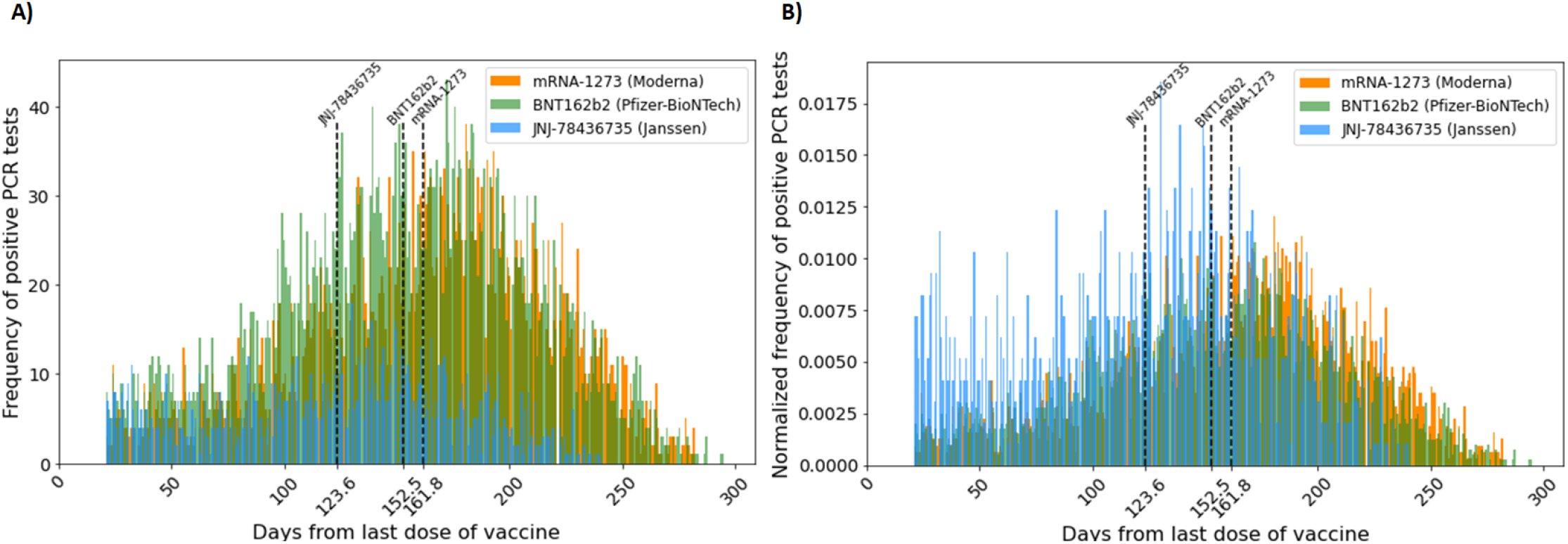
(A) Frequency of vaccine breakthrough infection (count per day) starting 21 days after the administration of the last COVID-19 vaccine dose. (B) Normalized frequency of vaccine breakthrough infection (define) starting 21 days after the administration of the last COVID-19 vaccine. Normalized frequency distributions of mRNA-1273 and BNT162b2 vaccine breakthrough cases are approximately distributed normally with the mean at 161.8 days and 152.8 days, respectively. That of JNJ-78436735 vaccine breakthrough cases is skewed to the right with the mean at 123.6 days. The lines represent the mean days until breakthrough for each vaccine.

**Figure 3.**
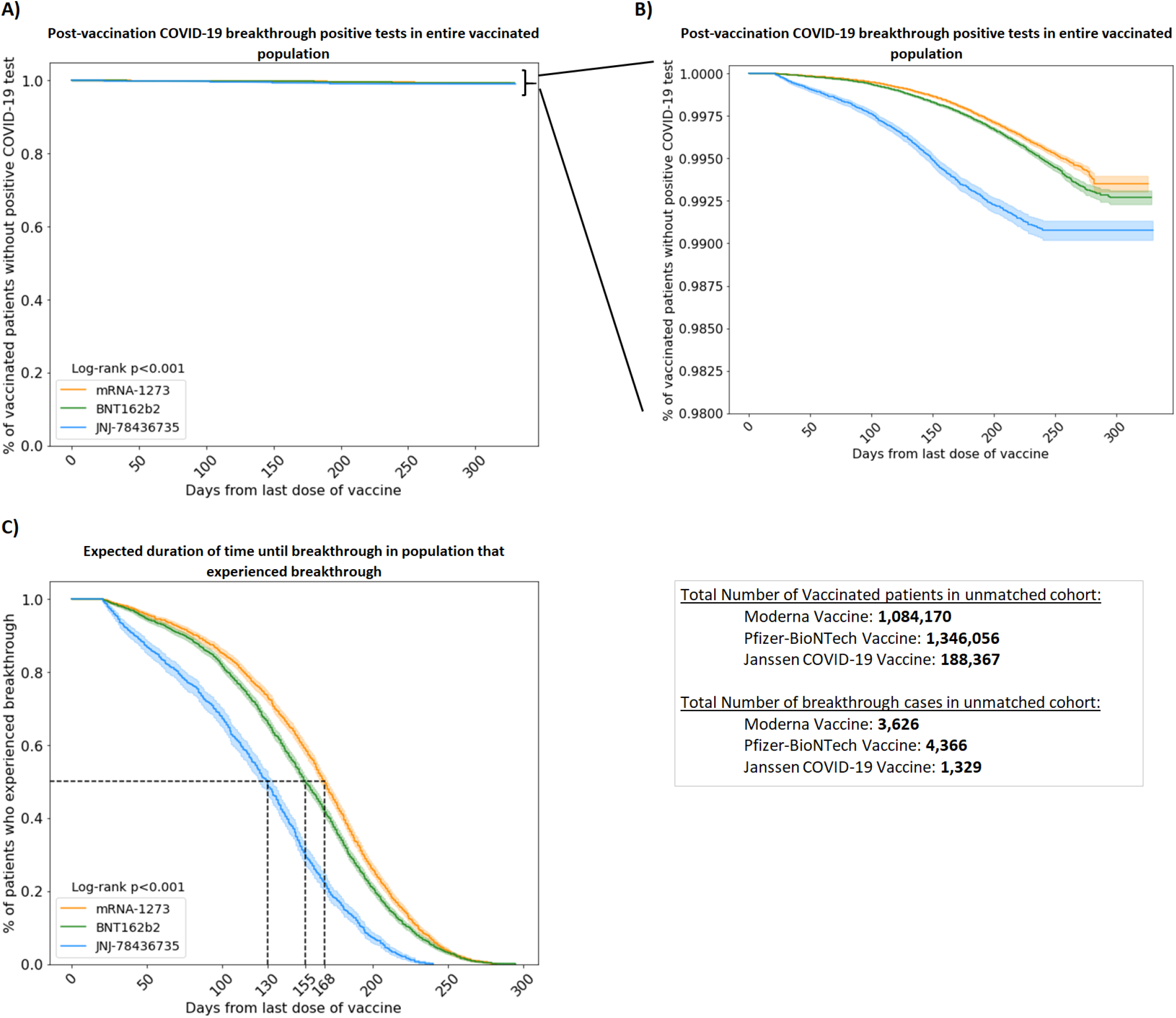
Vaccine immunity over time against SARS-CoV-2 infection for BNT162b2, mRNA-1273, and JNJ-78436735 vaccines. (A) Kaplan-Meier estimates for positive test result events based on the whole vaccinated population (including patients with positive test or negative test and not tested) (y-axis[0,1]), (B) Kaplan-Meier estimates for positive test result events based on the whole vaccinated population (including patients with positive test or negative test and not tested) (y-axis[0.98,1]), (C) Survival function of time to vaccine breakthrough using Kaplan-Meier estimate (Including only patients with positive test result). The lines represent the median days until breakthrough for each vaccine.

**Figure 4.**
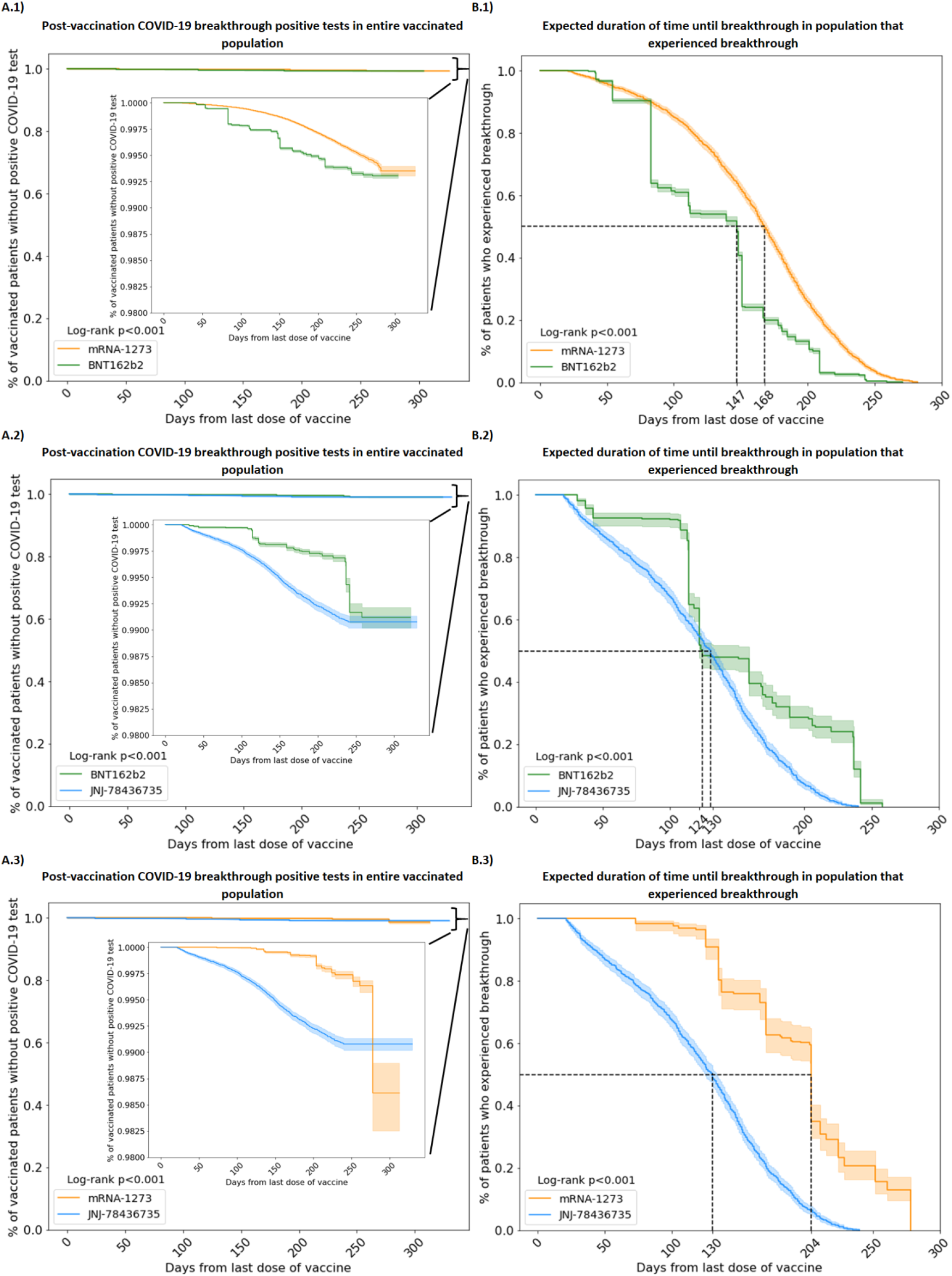
Matched cohort analysis for pairwise comparison of three administered vaccines, (A.1/B.1/C.1) mRNA-1273/BNT162b2, (A.2/B.2/C.2) BNT162b2/JNJ-78436735, and (A.3/B.3/C.3) mRNA-1273/JNJ-78436735. The lines represent the median days until breakthrough for each vaccine.

### Infection-induced immunity cohort

Descriptive analyses for the infection-induced immunity cohort are given in Supplementary Table 3. Of 191,722 patients who tested positive for COVID-19 during the study period, 64,424 patients were eligible as the infection-induced immunity cohort. The mean age of this cohort was 43.33 (± 18.63) years old. This cohort was enriched with 18-44 age group, female, non-Hispanic/Latino, White/Caucasian, and patients with invalid geographical information. This cohort was categorized into three groups based on the outcome; 567 (0.88%) patients in the reinfection subgroup, 11,067 (17.18%) in the positive-negative subgroup, and 52,507 (81.50%) in the positive-not tested subgroup. Similar to the vaccine-induced immunity breakthrough subgroup, patients between the ages of 18-44 had the highest rate of reinfection; however, they made up a larger fraction: 55.73% vs. 25.6%. The reinfection group was enriched with 18-44 age group, female, non-Hispanic/Latino, White/Caucasian, and California residents.

Patients who died within 90 days of their initial infection were excluded. Patients who died after 90 days were considered unlikely to still be infected from their initial case and were right-censored. The estimated survival functions (Figure 5A and 5B) for this cohort displays an initial exponential decay followed by a linear decrease. The distribution of times itself also displayed approximately exponential behavior which asymptotically approaches a uniform distribution (Figure 5D). However, we only displayed cases with reinfection times up to 350 days to compare to the vaccine-induced immunity cohort. About 5% of the total cases fall between 350 and 600 days and are not displayed here but are included in the overall statistics and survival analysis. The inclusion of cases beyond 350 days gives more pronounced exponential behavior for both the distribution and the survival functions. Additionally, if we relax the assumption that reinfection occurs >90 days, we also see a more pronounced exponential distribution and survival function, which drastically reduces the mean and median reinfection times (Supplementary Figure 1). We find that the probability of avoiding reinfection is 0.995 at 180 days, 0.989 at 350 days and 0.987 at 600 days. However, this is only applicable for patients who survived their initial infection. Adjusting for the mortality rate, these probabilities become 0.945 at 180 days, 0.939 at 350 days, and 0.937 at 600 days.

**Figure 5.**
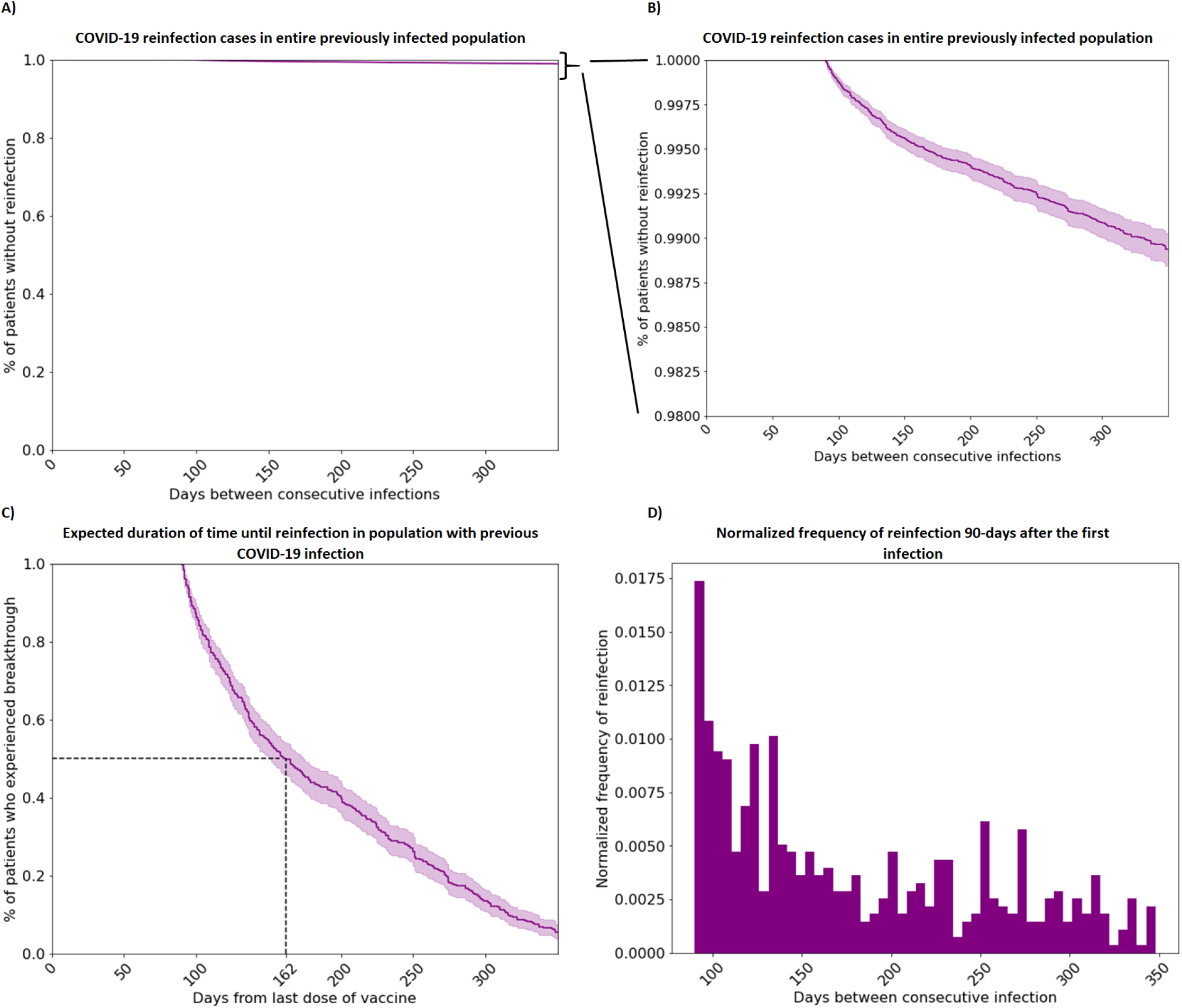
(A) Kaplan-Meier estimates for the survival function for infection-induced immunity. Reinfection is defined by a positive COVID-19 PCR or NAAT test 90 days after the previous positive test. Range for the X axis extends to 600+ days, but only 350 days is displayed for controlled comparison with the vaccinated cohort (y-axis[0,1]). (B) More detailed close-up view of A, (y-axis[0.98,1]) (C) Kaplan-Meier estimates for the survival function restricted to patients who have been reinfected. ∼5% of reinfected patients had more than 350 days between infections. (D) Normalized frequency of reinfection 9 months after the first infection.

## Discussion

In this study, we examined the acquired immunity through three vaccines (mRNA-1273, BNT162b2, JNJ-78436735) that have been administered in the United States by characterizing breakthrough cases in a large, vaccinated population across California, Oregon, Washington, Alaska, and Montana. We also conducted pairwise-comparison of the acquired immunity of the vaccines, matching individual vaccine cohorts based on sex, ethnicity, age, and timing of the completion of the recommended initial dose(s). As of November 5th, 2021, 2,627,914 patients (mRNA-1273: 1,087,796; BNT162b2: 1,350,422; JNJ-78436735: 189,696) were fully vaccinated and 0.36% (N=9,321) tested positive after vaccination. All three vaccines showed a high probability of survival against breakthrough cases (mRNA-1273: 0.997, BNT162b2: 0.997, JNJ-78436735: 0.992 in 180 days). In terms of individual performance, two doses of mRNA-1273 was the most effective and a single dose of JNJ-78436735 was least effective among the three administered immunizations (168, 155, and 130 median days to breakthrough, P<0.05). These results remained robust in propensity score analyses. The present results support evidence of previous studies (9–11).

Additionally, we examined the acquired immunity through prior COVID-19 infection for preventing reinfection. We found a median survival time of 162 days and a mean of 191 days, with an asymptotic probability for avoiding reinfection of .995 at 180 days, which is comparable to vaccine-induced immunity breakthrough. We must be careful in comparing these numbers since we have a larger threshold between reinfected cases and breakthroughs. There is also a substantially greater risk of mortality subsequent to SARS-CoV-2 infection than there is to vaccination. We adjust for this by multiplying the resulting survival probability for reinfection by the probability of surviving after 90 days. This gives an adjusted survival of reinfection probability of .945 at 180 days.

Of note, people aged 18-44 years make up a significantly larger portion of the reinfected cohort than the breakthrough cohort (55.7% vs. 25.6%). This demographic also made up over half of the total workforce in the US in 2020, which could contribute to increased exposure. Another interesting difference between the demographics is the incidence of comorbidities. There seems to be a larger proportion of patients with comorbidities in the vaccinated cohort than in the reinfected cohort. Because chronic comorbidities have been reported as a risk factor for more severe COVID-19, patients with these comorbidities may have been more likely than the general population to get vaccinated. Further, we found that the incidence rate of reinfection was higher in the infection-induced immunity cohort than that of the vaccine-induced cohort (0.9% vs. 0.4% of the eligible populations of the infection-induced immunity and vaccine-induced immunity cohorts, respectively). This finding is smaller than that reported in a previous study (21), which suggested that the vaccine-induced immunity population had a six-fold higher risk of COVID-19 infection than COVID-induced immunity population. This study was different from our study as it was conducted in Israel and BNT162b2 was the only vaccine administered. Additionally, genomic mutations of COVID-19 (22) are distributed differently according to geographic location and pandemic time window.

Interestingly, the shape of the survival curve for infection-induced immunity contains no inflection point (Figure 3), unlike the survival curve for breakthrough infection (Figure 4). Since these curves are cumulative distributions as a function of time, inflection points are extreme in the probability distributions. For breakthroughs, the probability distributions are approximately normal, leading to a maximum, and therefore inflection point, around the mean whereas the distribution for infection-induced immunity is approximately exponential, which indicates that the probability of reinfection is highest towards the start of the interval. Exponential distributions possess a property called “memorylessness”. Formally, if *T* is survival time, this means *Pr*(*T* > *t* + *a*|*T* > *a*) = *Pr*(*T* > *t*), implying that the probability that one will get reinfected within a fixed number of days will always be the same. This is not applicable for vaccine-induced immunity, where distribution is approximately normal, leading to a complementary error function for its survival function. In general, it is difficult to say if *Pr*(*T* > *t* + *a*|*T* > *a*) is increasing or decreasing as a function of *a*.

Another interesting aspect of these distributions is the tails. Probability distributions must have a total measure of 1 and tend to zero at infinity. This is slightly counterintuitive in the current context, since one would expect the probability of getting reinfected would increase over time due to waning immunity. This could be attributed to i) lack of data over an extended period of time and ii) lack of access to positive tests outside of PSJH. i) comes from the fact that a majority of vaccinated individuals got their vaccine in the last year and may still get infected, especially as new variants spread through the population. As time progresses, we would expect the mean and median times to increase as patients in our cohort who have not been infected, experience breakthroughs. For ii), home and rapid tests make up a significant portion of all COVID-19 tests taken and are not accessible through the PSJH. Still, the actual asymptotic survival probabilities are very high.

### Strengths and Limitations

In this study, we characterized both vaccine-induced immunity cohort and infection-induced immunity cohort. Direct comparison between vaccine-induced and infection-induced immunity cohorts is unreasonable because it is difficult to establish the time that the infection-induced immunity cohort recovered from COVID-19 and gained immunity. We instead conducted parallel analyses between vaccine-induced and infection-induced immunity cohorts.

We analyzed a large sample that represents the general population of the U.S. west coast. Previous published studies in the U.S. (9,23–27) are limited to a cohort with specific characteristics, such as health-care workers (6,15,23,27), patients with specific comorbidities (28–30), veterans (9,24,29,31), patients of academic hospitals (32,33). PSJH is a community-served hospital that covers five states of the west coast in both rural and urban areas. As we observed consistent results regarding vaccine-induced immunity (mRNA-1273 > BNT162b2 > JNJ-78436735) from previous studies (9,10,15) conducted in a cohort with special characteristics, our result adds validity and increases generalizability of previous studies. To our knowledge, our study had the longest observed duration for time to breakthrough/reinfection, encompassing both waning vaccine- and infection-induced immunity (21,34,35) and evolving variants (36).

We performed propensity score matching analysis pairwise between individual vaccines. We had a total of 2,627,914 patients (mRNA-1273: 1,087,796; BNT162b2: 1,350,422; JNJ-78436735: 189,696). We matched on sex, age, ethnicity, geographical location, and administered time of the last dose of vaccination. Geographical location and the time of the last dose of vaccination were proxy for environmental exposures. We adjusted for time to address how vaccination eligibility has changed over time based on the type of job. High-risk critical workers were eligible for vaccination in the first quarter of 2021 (37–42). People who were able to remotely work were eligible in the second quarter of 2021 (37,39–43). Patients were also matched by state of residence, in consideration of geographic differences in policies and vaccination percentages.

A potential weakness of this study is that, although PSJH captures vaccination records through bidirectional linkage state immunization registries, it does not necessarily have results for COVID-19 tests from outside of the PSJH system. Future analyses would benefit from requiring access to this data from these alternative data sources, as well as sequencing, titer level and other deep immunophenotyping data (44,45). Two other limitations are that variant types are infrequently tracked in the EHR, and we cannot precisely determine whether reinfection defined in this study is an actual reinfection, as opposed to an unusually long lingering initial infection. However, we chose a conservative threshold, 90 days after the first positive test, to determine the reinfection cases. Studies to date suggest that the maximum duration of SARS-CoV-2 RNA detection in upper respiratory tract specimens is 12 weeks (84 days) (46,47) after symptom onset and reinfection does not occur within 90 days of first COVID-19 infection or illness (48). Moreover, sensitivity analysis which included patients with reinfection times in between 21 days and 90 days drastically reduced the median reinfection time to 37 days, invalid to be considered as reinfection, and the asymptotic survival probability from 0.987 to 0.952 (Supplementary Figure 1). As a note, our choice to KNN function with replacement to match the propensity scores improves the average quality of matching, but because controls are used multiple times, the control group contains less information.

Additionally, our data is limited due to incongruencies in vaccine rollout between the three vaccines. Initially, only healthcare and other essential workers had access to the vaccine, with redistribution of the larger public coming only later. This means that a majority of patients who received the vaccine have an upper bound on the possible times they can be considered for reinfection. As time progresses, these people may experience breakthroughs at later times which would skew the distribution to the right, increasing the mean and median times reported. As such, the numbers reported in Figures 2, 3, 4 and 5 may increase in time as more people are eligible to get infected at later times.

## Conclusion

Here we presented demographic information, survival functions and probability distributions for patients experiencing vaccination breakthrough or post-infection reinfection within the PSJH network, showing that all three vaccines (BNT162b2, mRNA-1273, and JNJ-78436735) have very low risk of breakthrough in the real world over 350 days. We also observed infection-induced immunity, though we did not make a direct comparison with vaccine-induced immunity for the reasons discussed above. However, the risks associated with COVID-19 infection are far, far greater than any marginal advantages acquired by prior infection.

## Data Availability

All data produced in the present study are available upon reasonable request to the authors

## Conflict of Interest Disclosures

None of the authors have a conflict of interest with this study.

## Funding/Support

This work was funded in part by the Swedish Medical Center Foundation.

## Acknowledgement

We are grateful to Providence St. Joseph Health for sharing their data engineering expertise and computational resources. We would also like to acknowledge SNOMED International for developing and maintaining SNOMED-CT.

## Author contributions

JJH, VD, AMB, YH, SM conceptualized the study. SM, AMB were involved in the EHR data extraction, data cleaning, and codification. SM, AMB, YH performed data analysis including statistical and survival analysis. JJH supervised implementation and provided administrative and material support. AMB, YH, SM prepared the manuscript with critical revision of the manuscript for important intellectual content provided by JJH and JDG. All authors reviewed and approved the final version of the manuscript.

## Notes

### Competing Interest Statement

The authors have declared no competing interest.

### Author Declarations

Ethics committee of Institute for Systems Biology gave ethical approval for this work.

## References

1. Roser M, Ritchie H, Ortiz-Ospina E, Hasell J. Coronavirus pandemic (COVID-19). Our world in data. 2020;

2. National Center for Immunization and Respiratory Diseases (NCIRD), Division of Viral Diseases. Science Brief: COVID-19 Vaccines and Vaccination. In: CDC COVID-19 Science Briefs. Atlanta (GA): Centers for Disease Control and Prevention (US); 2021.

3. Griffin JB, Haddix M, Danza P, Fisher R, Koo TH, Traub E, et al. SARS-CoV-2 Infections and Hospitalizations Among Persons Aged ≥16 Years, by Vaccination Status - Los Angeles County, California, May 1-July 25, 2021. MMWR Morb Mortal Wkly Rep. 2021 Aug 27;70(34):1170–6.

4. Rosenberg ES, Holtgrave DR, Dorabawila V, Conroy M, Greene D, Lutterloh E, et al. New COVID-19 Cases and Hospitalizations Among Adults, by Vaccination Status - New York, May 3-July 25, 2021. MMWR Morb Mortal Wkly Rep. 2021 Sep 17;70(37):1306–11.

5. Nanduri S, Pilishvili T, Derado G, Soe MM, Dollard P, Wu H, et al. Effectiveness of Pfizer-BioNTech and Moderna Vaccines in Preventing SARS-CoV-2 Infection Among Nursing Home Residents Before and During Widespread Circulation of the SARS-CoV-2 B.1.617.2 (Delta) Variant - National Healthcare Safety Network, March 1-August 1, 2021. MMWR Morb Mortal Wkly Rep. 2021 Aug 27;70(34):1163–6.

6. Fowlkes A, Gaglani M, Groover K, Thiese MS, Tyner H, Ellingson K, et al. Effectiveness of COVID-19 Vaccines in Preventing SARS-CoV-2 Infection Among Frontline Workers Before and During B.1.617.2 (Delta) Variant Predominance - Eight U.S. Locations, December 2020-August 2021. MMWR Morb Mortal Wkly Rep. 2021 Aug 27;70(34):1167–9.

7. Dan JM, Mateus J, Kato Y, Hastie KM, Yu ED, Faliti CE, et al. Immunological memory to SARS-CoV-2 assessed for up to 8 months after infection. Science [Internet]. 2021 Feb 5;371(6529). Available from: http://dx.doi.org/10.1126/science.abf4063

8. Organization WH, Others. COVID-19 natural immunity: scientific brief, 10 May 2021 [Internet]. World Health Organization; 2021. Available from: https://apps.who.int/iris/bitstream/handle/10665/341241/WHO-2019-nCoV-Sci-Brief-Natural-immunity-2021.1-eng.pdf

9. Cohn BA, Cirillo PM, Murphy CC, Krigbaum NY, Wallace AW. SARS-CoV-2 vaccine protection and deaths among US veterans during 2021. Science. 2021 Nov 4;eabm0620.

10. Self WH, Tenforde MW, Rhoads JP, Gaglani M, Ginde AA, Douin DJ, et al. Comparative Effectiveness of Moderna, Pfizer-BioNTech, and Janssen (Johnson & Johnson) Vaccines in Preventing COVID-19 Hospitalizations Among Adults Without Immunocompromising Conditions - United States, March-August 2021. MMWR Morb Mortal Wkly Rep. 2021 Sep 24;70(38):1337–43.

11. Grannis SJ, Rowley EA, Ong TC, Stenehjem E, Klein NP, DeSilva MB, et al. Interim Estimates of COVID-19 Vaccine Effectiveness Against COVID-19-Associated Emergency Department or Urgent Care Clinic Encounters and Hospitalizations Among Adults During SARS-CoV-2 B.1.617.2 (Delta) Variant Predominance - Nine States, June-August 2021. MMWR Morb Mortal Wkly Rep. 2021 Sep 17;70(37):1291–3.

12. Tartof SY, Slezak JM, Fischer H, Hong V, Ackerson BK, Ranasinghe ON, et al. Effectiveness of mRNA BNT162b2 COVID-19 vaccine up to 6 months in a large integrated health system in the USA: a retrospective cohort study. Lancet. 2021 Oct 16;398(10309):1407–16.

13. Dagan N, Barda N, Kepten E, Miron O, Perchik S, Katz MA, et al. BNT162b2 mRNA Covid-19 Vaccine in a Nationwide Mass Vaccination Setting. N Engl J Med. 2021 Apr 15;384(15):1412–23.

14. Thomas SJ, Moreira ED Jr, Kitchin N, Absalon J, Gurtman A, Lockhart S, et al. Safety and Efficacy of the BNT162b2 mRNA Covid-19 Vaccine through 6 Months. N Engl J Med. 2021 Nov 4;385(19):1761–73.

15. Pilishvili T, Gierke R, Fleming-Dutra KE, Farrar JL, Mohr NM, Talan DA, et al. Effectiveness of mRNA Covid-19 Vaccine among U.S. Health Care Personnel. N Engl J Med. 2021 Dec 16;385(25):e90.

16. CDC. Investigative Criteria for Suspected Cases of SARS-CoV-2 Reinfection (ICR) [Internet]. 2021 [cited 2021 Dec 13]. Available from: https://www.cdc.gov/coronavirus/2019-ncov/php/invest-criteria.html

17. Sheehan MM, Reddy AJ, Rothberg MB. Reinfection Rates Among Patients Who Previously Tested Positive for Coronavirus Disease 2019: A Retrospective Cohort Study. Clin Infect Dis. 2021 Nov 16;73(10):1882–6.

18. Rosenbaum PR, Rubin DB. The Central Role of the Propensity Score in Observational Studies for Causal Effects. Biometrika. 1983;70(1):41–55.

19. Harder VS, Stuart EA, Anthony JC. Propensity score techniques and the assessment of measured covariate balance to test causal associations in psychological research. Psychol Methods. 2010 Sep;15(3):234–49.

20. Pedregosa F. Scikit-learn: Machine Learning in Python. J Mach Learn Res. 2011;12:2825–30.

21. Gazit S, Shlezinger R, Perez G, Lotan R, Peretz A, Ben-Tov A, et al. Comparing SARS-CoV-2 natural immunity to vaccine-induced immunity: reinfections versus breakthrough infections [Internet]. bioRxiv. 2021. Available from: http://medrxiv.org/lookup/doi/10.1101/2021.08.24.21262415

22. Mercatelli D, Giorgi FM. Geographic and Genomic Distribution of SARS-CoV-2 Mutations. Front Microbiol. 2020 Jul 22;11:1800.

23. Bergwerk M, Gonen T, Lustig Y, Amit S, Lipsitch M, Cohen C, et al. Covid-19 Breakthrough Infections in Vaccinated Health Care Workers. N Engl J Med. 2021 Oct 14;385(16):1474–84.

24. Dickerman BA, Gerlovin H, Madenci AL, Kurgansky KE, Ferolito BR, Figueroa Muñiz MJ, et al. Comparative Effectiveness of BNT162b2 and mRNA-1273 Vaccines in U.S. Veterans. N Engl J Med [Internet]. 2021 Dec 1; Available from: https://doi.org/10.1056/NEJMoa2115463

25. Cohn BA, Cirillo PM, Murphy CC, Krigbaum NY, Wallace AW. Breakthrough SARS-CoV-2 infections in 620,000 U.S. Veterans, February 1, 2021 to August 13, 2021 [Internet]. bioRxiv. 2021. Available from: http://medrxiv.org/lookup/doi/10.1101/2021.10.13.21264966

26. Ramirez E, Wilkes RP, Carpi G, Dorman J, Bowen C, Smith L. SARS-CoV-2 breakthrough infections in fully vaccinated individuals [Internet]. bioRxiv. medRxiv; 2021. Available from: http://medrxiv.org/lookup/doi/10.1101/2021.06.21.21258990

27. Shamier MC, Tostmann A, Bogers S, de Wilde J, IJpelaar J, van der Kleij WA, et al. Virological characteristics of SARS-CoV-2 vaccine breakthrough infections in health care workers [Internet]. bioRxiv. 2021. Available from: http://medrxiv.org/lookup/doi/10.1101/2021.08.20.21262158

28. Qin CX, Moore LW, Anjan S, Rahamimov R, Sifri CD, Ali NM, et al. Risk of Breakthrough SARS-CoV-2 Infections in Adult Transplant Recipients. Transplantation. 2021 Nov 1;105(11):e265–6.

29. Sankary KM, Sippel JL, Eberhart AC, Burns SP. Breakthrough cases of COVID-19 in vaccinated United States Veterans with spinal cord injuries and disorders. Spinal Cord. 2021 Oct;59(10):1132–3.

30. Di Fusco M, Moran MM, Cane A, Curcio D, Khan F, Malhotra D, et al. Evaluation of COVID-19 vaccine breakthrough infections among immunocompromised patients fully vaccinated with BNT162b2. J Med Econ. 2021 Jan;24(1):1248–60.

31. Sharma A, Oda G, Holodniy M. COVID-19 vaccine breakthrough infections in Veterans Health Administration [Internet]. bioRxiv. 2021. Available from: http://medrxiv.org/lookup/doi/10.1101/2021.09.23.21263864

32. Servellita V, Sotomayor-Gonzalez A, Gliwa AS, Torres E, Brazer N, Zhou A, et al. Predominance of antibody-resistant SARS-CoV-2 variants in vaccine breakthrough cases from the San Francisco Bay Area, California [Internet]. bioRxiv. 2021. Available from: http://medrxiv.org/lookup/doi/10.1101/2021.08.19.21262139

33. Duerr R, Dimartino D, Marier C, Zappile P, Levine S, François F, et al. Clinical and genomic signatures of rising SARS-CoV-2 Delta breakthrough infections in New York. medRxiv [Internet]. 2021 Dec 8; Available from: http://dx.doi.org/10.1101/2021.12.07.21267431

34. Post N, Eddy D, Huntley C, van Schalkwyk MCI, Shrotri M, Leeman D, et al. Antibody response to SARS-CoV-2 infection in humans: A systematic review. PLoS One. 2020 Dec 31;15(12):e0244126.

35. Peluso MJ, Takahashi S, Hakim J, Kelly JD, Torres L, Iyer NS, et al. SARS-CoV-2 antibody magnitude and detectability are driven by disease severity, timing, and assay. Sci Adv [Internet]. 2021 Jul;7(31). Available from: http://dx.doi.org/10.1126/sciadv.abh3409

36. CDC. SARS-CoV-2 Variant Classifications and Definitions [Internet]. 2021 [cited 2021 Dec 22]. Available from: https://www.cdc.gov/coronavirus/2019-ncov/variants/variant-classifications.html

37. COVID-19 Vaccine Eligibility - Phases and Eligibility [Internet]. San Diego County. [cited 2021 Dec 22]. Available from: https://www.sandiegocounty.gov/content/sdc/hhsa/programs/phs/community_epidemiology/dc/2019-nCoV/vaccines/phases.html

38. Who goes next: Washington releases next phase of vaccine prioritization [Internet]. [cited 2021 Dec 17]. Available from: https://www.doh.wa.gov/Newsroom/Articles/ID/2554/Who-goes-next-Washington-releases-next-phase-of-vaccine-prioritization

39. Dooling K, Marin M, Wallace M, McClung N, Chamberland M, Lee GM, et al. The Advisory Committee on Immunization Practices’ Updated Interim Recommendation for Allocation of COVID-19 Vaccine - United States, December 2020. MMWR Morb Mortal Wkly Rep. 2021 Jan 1;69(5152):1657–60.

40. Alaska Department of Health and Social Services. COVID vaccine Alaska Allocation Guidelines [Internet]. Alaska Department of Health and Social Services; 2021 Mar. Available from: https://dhss.alaska.gov/dph/Epi/id/SiteAssets/Pages/HumanCoV/COVIDvaccine_AlaskaAllocationGuidelines.pdf

41. Montana Department of Public Health and Human Services. Montana COVID-19 Vax Plan [Internet]. Montana Department of Public Health and Human Services ; January, 5 2021. Available from: https://dphhs.mt.gov/assets/Coronavirus/MontanaCOVID-19VaxPlanV5.pdf

42. Oregon Health Authority. COVID-19-Vaccination-Plan-Oregon [Internet]. Oregon Health Authority; 2020 Dec. Available from: https://www.oregon.gov/oha/covid19/Documents/COVID-19-Vaccination-Plan-Oregon.pdf

43. COVID-19 vaccine distribution update from the Washington State Department of Health [Internet]. [cited 2021 Dec 17]. Available from: https://www.doh.wa.gov/Newsroom/Articles/ID/2724/COVID-19-vaccine-distribution-update-from-the-Washington-State-Department-of-Health

44. Goldman JD, Wang K, Roltgen K, Nielsen SCA, Roach JC, Naccache SN, et al. Reinfection with SARS-CoV-2 and Failure of Humoral Immunity: a case report. medRxiv [Internet]. 2020 Sep 25; Available from: http://dx.doi.org/10.1101/2020.09.22.20192443

45. Seow J, Graham C, Merrick B, Acors S, Pickering S, Steel KJA, et al. Longitudinal observation and decline of neutralizing antibody responses in the three months following SARS-CoV-2 infection in humans. Nature Microbiology. 2020 Oct 26;5(12):1598–607.

46. Li N, Wang X, Lv T. Prolonged SARS-CoV-2 RNA shedding: Not a rare phenomenon. J Med Virol. 2020 Nov;92(11):2286–7.

47. Liu W-D, Chang S-Y, Wang J-T, Tsai M-J, Hung C-C, Hsu C-L, et al. Prolonged virus shedding even after seroconversion in a patient with COVID-19. J Infect. 2020 Aug;81(2):318–56.

48. CDC. Ending Isolation and Precautions for People with COVID-19: Interim Guidance [Internet]. 2021 [cited 2021 Dec 22]. Available from: https://www.cdc.gov/coronavirus/2019-ncov/hcp/duration-isolation.html

